# Nonparametric serial interval estimation

**DOI:** 10.1101/2024.10.16.24315600

**Authors:** Oswaldo Gressani, Niel Hens

## Abstract

The serial interval of an infectious disease is a key instrument to understand transmission dynamics. Estimation of the serial interval distribution from illness onset data extracted from transmission pairs is challenging due to the presence of censoring and state-of-the-art frequentist or Bayesian methods mostly rely on parametric models. We present a fully data-driven methodology to estimate the serial interval distribution based on (coarse) serial interval data. The proposal combines a nonparametric estimator of the cumulative distribution function with the bootstrap and yields point and interval estimates of any desired feature of the serial interval distribution. Algorithms underlying our approach are simple, fast and stable, and are thus easily implementable in any programming language most desired by modelers from the infectious disease community. The nonparametric routines are included in the EpiLPS package for ease of implementation. Our method complements existing parametric approaches for serial interval estimation and permits to straightforwardly analyze past, current, or future illness onset data streams.

## 1 Introduction

The serial interval (SI) of an infectious disease is an epidemiological delay characterizing a duration between two well-defined events related to a disease. It represents the time between symptom(s) onset in a primary case or infector and the symptom(s) onset in a secondary case or infectee (Simpson, 1948). This time delay can be negative as nothing restrains the illness onset time of the infector to be larger than the onset time of the infectee. In the literature, this interval is also known as the clinical onset serial interval (Cowling et al., 2009; Te Beest et al., 2014). A different, but closely related delay quantity is the generation interval (GI), which is defined as the duration between infection events in an infector-infectee pair (Svensson, 2007). Contrary to the SI, the GI is a delay quantity that is always positive. The timing of an infection event is typically less likely to be observed than the timing of a symptom event and it is common practice to approximate the distribution of generation times by the SI distribution (Lehtinen et al., 2021; Chen et al., 2022). Serving as a proxy for generation intervals, serial intervals can be used as an instrument to measure the time scale of disease transmission (Park et al., 2021) and are therefore key in linking the epidemic growth rate with the time-varying reproduction number (Wallinga and Lipsitch, 2007; Torneri et al., 2021). The crucial role played by serial intervals in disease transmission models emphasizes the need to have reliable, stable, and replicable statistical methodologies to estimate this transmission interval. Ideally, these methodologies should also follow best practices as recently described in Charniga et al. (2024).

Different methods exist to estimate the distribution and features of the serial interval of an infectious disease based on data. When time intervals of illness onset between infectors and infectees are observed, the data is considered as a random sample from the population. In that case, essential features of the serial interval are estimated by either directly computing summary statistics from empirical serial intervals (e.g. mean, median, standard deviation) or by fitting a parametric distribution to observed data (Bóelle et al., 2011; Griffin et al., 2020). Parametric methods are by far the most common and usually include the Lognormal, Weibull, Gamma or Gaussian distributions (Lessler et al., 2009; Cowling et al., 2010; Li et al., 2020; Nishiura et al., 2020; Ma et al., 2020; Kremer et al., 2022). Estimation of model parameters is typically carried out with the maximum likelihood principle or by using the Bayesian approach, and often on a few observations. To our knowledge, only few attempts have been made in applying nonparametric methods to serial interval data analysis. For instance, Cowling et al. (2009) compute a nonparametric estimate of the cumulative distribution function of the serial interval of influenza based on the method of Turnbull (1976) to see whether different parametric models are in agreement with it; and Mettler et al. (2020) use the nonparametric bootstrap to compute confidence intervals for the clinical onset SI of SARS-CoV-2.

By definition, serial intervals involve transmission pairs. It means that a minimal requirement for SI estimation is to have data on symptom(s) onset times for the infector and infectee. Such data can be extracted from contact tracing programmes, which permit to gain knowledge about who infected whom and provide information on timings of symptoms in infector-infectee pairs (Yang et al., 2020; Müller and Kretzschmar, 2021). Commonly, serial interval data are coarse in that only lower and upper limits of illness onset timing is observed. This characteristic is known as censoring and adds a layer of complexity to the estimation problem. If coarseness concerns either the infector or infectee, data are said to be single interval-censored; and if coarseness affects both actors in the transmission pair, data are called doubly interval-censored (Reich et al., 2009). Thinking from a continuous time perspective, serial interval data is more often than not doubly interval-censored due to the time resolution of reporting. When the time resolution for reporting illness onset is a calendar day (as is often the case), coarseness is inherent to the calendar day, i.e. the precise timing of illness onset within the reported calendar day remains unknown. Therefore, even if exact calendar dates are observed, it is good practice to still consider the data as doubly interval-censored (Charniga et al., 2024).

Despite the large number of studies conducted on the serial interval of different pathogens, most methods are difficult or impossible to reproduce in the sense that independent researchers are confronted with serious difficulties in reusing existing procedures to new data (Gandrud, 2018). The field of infectious disease modeling suffers from alarmingly low computational reproducibility rates (Henderson et al., 2024), which hinders applicability and misaligns with pandemic preparedness objectives. This reproducibility conundrum has several causes. For instance, recent meta-epidemiological surveys found that very few publications share code or data (Collins and Alexander, 2022; Zavalis and Ioannidis, 2022). Other potential causes are code incompleteness and complex dependencies among multiple scripts without clear guidelines regarding computation order (Henderson et al., 2024). The study of Vink et al. (2014) highlights that finding evidence supporting frequently cited serial interval values in the literature is a challenging task.

Hopefully, more applicable tools and methods have recently emerged to estimate epidemiological delay distributions. Originally developed for estimation of incubation period distributions, the methodology of Reich et al. (2009) is available in an R software package (Reich et al., 2021) and associated routines are embedded in the EpiEstim package of Cori et al. (2013) to estimate the serial interval (Thompson et al., 2019). Vink et al. (2014) reanalyze published serial interval data on different respiratory infections by using a common statistical method and provide R code and data sets for reproducibility. The epidist R package (Park et al., 2024) is also operational for serial interval estimation and accounts for censoring and truncation. These tools rely on parametric methods imposing distributional assumptions on the serial interval distribution and leave no room for data-driven inference.

In an attempt to complement the above-mentioned parametric methods, we develop a nonparametric approach to estimate the serial interval distribution based on coarsely observed illness onset data. The proposed method is entirely data-driven and applicable on a wide range of serial interval data commonly analyzed in the literature. Its chief merits are its simplicity and the fact that it relies on two powerful statistical tools, namely the inverse-cdf method and the bootstrap. Since R is among the most popular programming languages used in the infectious disease modeling community (Batra et al., 2021; Henderson et al., 2024), the computer code underlying our nonparametric methodology is included in the EpiLPS package (Gressani, 2021; Gressani et al., 2022, 2024; Sumalinab et al., 2024) available on the Comprehensive R Archive Network (CRAN) repository. Source code comes in a lightweight format and spans only a few lines. It can thus be easily translated in another programming language if needed (e.g. Python).

Next, we present our nonparametric estimator and briefly discuss some of its theoretical properties. The performance of our method is assessed in extensive simulation scenarios. Applications to transmission pair data extracted from previous analyses for a diverse set of pathogens underlines the wide, general, and straightforward applicability of our approach. The article concludes with a discussion on the main strengths and limitations of the proposed nonparametric toolbox for serial interval estimation.

## 2 Methods

### 2.1 Notation

Let 𝒮 be a real-valued random variable representing the serial interval of an infectious disease and denote by *F*_*S*_ (*·*) the cumulative distribution function (cdf) of 𝒮 with *F*_*S*_ (*s*) = *P* (*S* ≤ *s*) ∀*s* ∈ ℝ. For the sake of generality, our model is formulated in continuous time. At the level of the *i*th transmission pair, 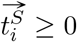 denotes the (finite) illness onset time of the infector and 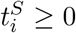 stands for the (finite) illness onset time of the infectee. In practice, the illness onset time is reported in calendar time and the serial interval is expressed in days. Conversion from calendar time to analysis time is usually done by assigning an integer to a calendar date. When illness onset timings are considered exactly observed, the serial interval for the *i*th transmission pair is simply 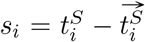. If the illness onset time of the infectee precedes the onset time of the infector, the serial interval is negative (*s*_*i*_ < 0) and the transmission event is called presymptomatic. In presence of coarse data, either 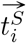 or 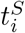 (single interval-censored data) or both (doubly interval-censored data) are only known to lie within a time interval, so that 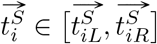, with 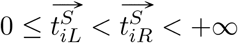 for the infector and 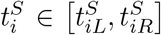, with 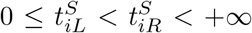 for the infectee. Following Reich et al. (2009), single or doubly interval-censored data can be transformed to interval-reduced data, containing the earliest possible and the latest possible serial interval time. For instance, if 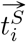 is exactly observed and the illness onset time of the infectee is interval-censored, the earliest possible SI time is 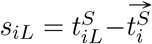 and the latest possible time is 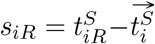. With doubly interval-censored observations, interval-reduced data is obtained by computing 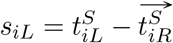 and 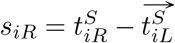. Note that both *s*_*iL*_ and *s*_*iR*_ can be negative and *s*_*iR*_ *> s*_*iL*_ will always hold. Even when 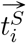 and 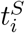 are exactly observed, we express the data as interval censored by constructing a displacement of length *δ* from *s*_*i*_ in both directions, i.e. *s*_*iL*_ = *s*_*i*_ − *δ* and *s*_*iR*_ = *s*_*i*_ +*δ* for an arbitrary *δ* (here *δ* = 0.5 to recover serial interval windows of unit length). For a sample containing *n* transmission pairs, the observed dataset has 2*n* elements and is denoted by 𝒟 = *{s*_1*L*_, *s*_1*R*_, …, *s*_*nL*_, *s*_*nR*_*}*. The set of features of 𝒮 is denoted by Θ_*S*_ = *{θ*_1_, *θ*_2_, …, *θ*_*J*_ *}* and contains all features of the serial interval that are of interest to the modeler. For example, if the aim is to estimate the mean, median and variance of 𝒮, the set Θ_*S*_ is a triplet with *θ*_1_ := 𝔼(𝒮), *θ*_2_ := inf*{s* ∈ ℝ : *F*_*S*_ (*s*) ≥ 0.5*}* and *θ*_3_ := 𝔼(𝒮 − 𝔼(𝒮))^2^. The goal is to provide data-driven point and interval estimates of elements of Θ_*S*_ based on 𝒟 without imposing any parametric assumption.

### 2.2 A simple nonparametric estimator of *F*_*S*_ (*·*)

Departing from a dataset *𝒟*, we build a nonparametric estimator of *F*_*S*_ (*·*) based on a simple idea. The available information at the level of the *i*th transmission pair is given by the left and right boundaries of the serial interval window, i.e. *s*_*iL*_ and *s*_*iR*_. Any point in the interval [*s*_*iL*_, *s*_*iR*_] corresponding to the true (and unobserved) serial interval of the *i*th pair can be seen as an observation from the continuous serial interval distribution (population). Therefore, the data points *s*_*iL*_ and *s*_*iR*_ extracted from the *i*th transmission pair can also be viewed as two observations or draws from *F*_*S*_ (*·*). A natural way to obtain a continuous estimate of *F*_*S*_ (*·*) based on *s*_*iL*_ and *s*_*iR*_ is to smooth the empirical cdf by linear interpolation (Bratley et al., 1987), yielding the following piecewise-linear empirical cdf (Kaczynski et al., 2012):

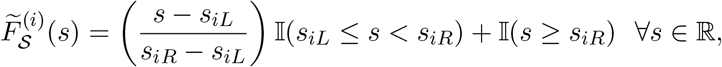

where 𝕀(*·*) is the indicator function. Extending this reasoning to the entire set of pairs in the dataset, we propose to estimate *F*_*S*_ (*·*) by averaging 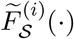 over all *n* transmission pairs. This data-driven estimator is given by:

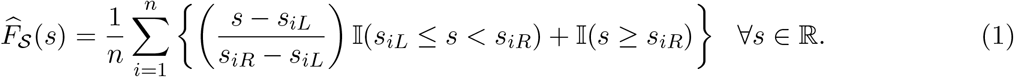

The above estimator is a (finite) linear combination of continuous functions 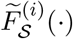 and is therefore itself a continuous function in ℝ. Moreover, it is a non-decreasing function since it essentially accumulates intervals when moving along the real line in the positive direction. It is also easy to verify that 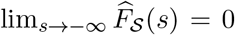 and 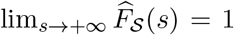, so that 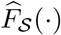 is a *bona fide* cdf. In addition, 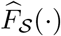 is a piecewise-linear function with breakpoints or “bends” arising at points in 𝒟, so that the cdf is almost everywhere differentiable (except at the set of points in 𝒟).

The properties of our estimator can be exploited to efficiently generate samples from 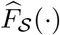, which can be viewed as approximate samples from the target serial interval distribution 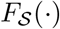. Let 𝒟_*O*_ = *{s*_(1)_, *s*_(2)_, …, *s*_(2*n*)_*}* denote the set of order statistics for 𝒟. Using the inverse-cdf method, the (pseudo) random-variate generation algorithm proceeds as follows (see e.g. Bratley et al., 1987):

- Generate a uniform random variable *U* in (0, 1), i.e. *U* ∼ 𝒰 (0, 1).
- Find *s*_(*i*)_ such that 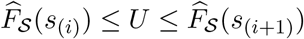.
- If 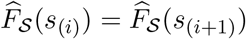 return *s*^*^ = *s*_(*i*)_.
- Else return 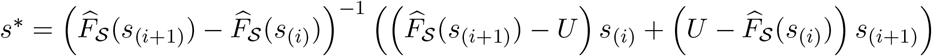.

### 2.3 The bootstrap

The bootstrap principle is used to calculate point and interval estimates of features of *S*. Using the random-variate generation algorithm presented in the previous section, *B* independent bootstrapsamples 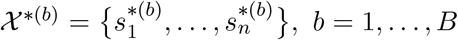 of size *n* are generated from 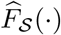. Let 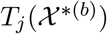 denote the statistic serving as an estimator of the feature *θ*_*j*_ ∈ Θ_*S*_. Based on the empirical bootstrap distribution 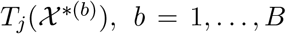, a point estimate of *θ*_*j*_ is given by the mean of the statistics generated by the resampling scheme, i.e. 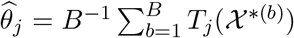. A 100(1 − *α*)% confidence interval for *θ*_*j*_ is given by 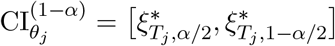, where 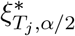 denotes the *α/*2 quantile and 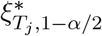 the 1 − *α/*2 quantile of the empirical bootstrap distribution. Most software has readily available routines to compute these quantiles (e.g. the *quantile* function in R).

## 3 Simulations

### 3.1 Data generating mechanism for artificial serial interval data

To simulate artificial serial interval data, we assume that the target SI distribution belongs to a parametric family indexed by a vector ***η*** and denote this by 𝒮∼ 𝒫_***η***_. In particular, we consider two distributions inspired from the literature. The first is a Gaussian distribution 𝒮 ∼ 𝒩 (2.8, 2.5^2^) with a mean of 2.8 days and a standard deviation of 2.5 days, mimicking the SI distribution of SARS-CoV-2 Omicron (Kremer et al., 2022), designated by the World Health Organization as a variant of concern (World Health Organization). The second is a Weibull distribution 𝒮∼ 𝒲 (2.36, 3.18) with shape 2.36 and scale 3.18 that imitates the SI distribution of influenza A (Lessler et al., 2009).

We denote by *A* the artificial serial interval dataset produced by our data generating mechanism (DGM). The DGM proceeds in a loop, where each iteration generates serial interval data for the *i*th transmission pair. At iteration *i*, a SI value is drawn from the target serial interval distribution *s*_*i*_ ∼ 𝒫_***η***_. Next, an arbitrary positive real number is assigned to the illness onset time of the infector through the uniform distribution 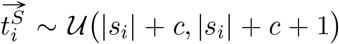, where *c >* 0 is a scalar that will be clarified later on. The illness onset time of the infectee is simply 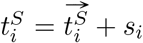 and note that the constraints 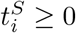 and 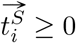 are satisfied.

In presence of censoring, a rule is needed to determine the left and right boundaries of the illness onset interval reported by a member of the *i*th transmission pair. We illustrate this rule for the infectee. The same rule holds for the infector. It is important to generate illness onset intervals of various widths 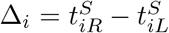 to mimic the fact that different individuals will typically report different illness onset intervals (heterogeneity of the population in the reporting process). In that direction, we assume that Δ_*i*_ is random and governed by a Gamma distribution Δ_*i*_ ∼ 𝒢(*a, b*) with shape *a* and rate *b*, so that 𝔼(Δ_*i*_) = *a/b* and 𝕍(Δ_*i*_) = *a/b*^2^. We fix *a* = *b* = 4, yielding illness onset intervals with an average width of one day. There is also less than 1% chance to generate interval widths above three days and roughly 95% chance to generate interval widths below two days.

Once Δ_*i*_ is available, another rule is required to determine the location of the boundaries 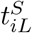 and 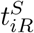 with respect to 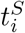. A simple rule would be to fix 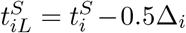 and 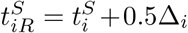. We believe that this rule is not realistic as it assumes that the individual is able to perfectly center the reported interval around the true illness onset time. A more realistic rule is to allow the interval to move randomly around 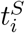. This can be achieved by generating a proportion according to a Beta distribution, say *ρ*_*i*_ ∼ *B*(5, 5), and interpret it as the proportion of the distance Δ_*i*_ that separates 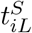 from 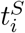. In other words, once *ρ*_*i*_ is obtained, simply compute 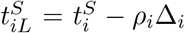 and 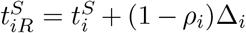, the left boundary of the interval is equal to 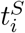 and 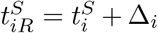 If *ρ*_*i*_ = 1, the right boundary of the interval is equal to 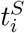 and the left boundary is 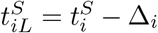 Any *ρ*_*i*_ ∈ (0, 1) generates a scenario in between these two extremes. The constant *c >* 0 used to generate 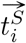 is simply there to ensure that 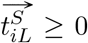 (and 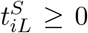). Without this constant, we could be in a scenario where 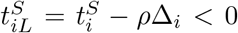. To avoid this, we fix *c* to an arbitrary large value (here *c* = 10). When data are exactly observed and after *n* cycles (reflecting a dataset with *n* transmission pairs), the DGM outputs *A* = *{s*_1_, …, *s*_*n*_*}*. In case of interval censoring, the DGM outputs 𝒜 = *{s*_1*L*_, *s*_1*R*_, …, *s*_*nL*_, *s*_*nR*_*}* after *n* cycles, where each element of 𝒜 contains either exactly observed data for at least one actor in the transmission pair (single interval censoring) or interval censored data for both actors in the transmission pair (doubly interval-censored data).

### 3.2 Simulation settings and results

The performance of our nonparametric method is assessed in different scenarios. For the Gaussian and Weibull target SI distributions described in Section 3.1, we consider small *n* = 10, medium *n* ∈ *{*20, 50*}* and large *n* = 100 sample sizes for doubly and single interval-censored data, as well as for exactly observed serial interval data (no censoring). This yields a total of 2 *×* 4 *×* 3 = 24 scenarios, which are summarized in Table 1. Each scenario involves *M* = 500 dataset replications and the performance of our approach is assessed on seven features of the serial interval *S*, namely, the mean, standard deviation (SD) and the 5th, 25th, 50th, 75th and 95th percentiles denoted by *q*0.05, *q*0.25, *q*0.50, *q*0.75 and *q*0.95, respectively. For each feature, we report the bias, empirical standard error (ESE) and root mean square error (RMSE), as well as the coverage probability (CP) and median interval width (ΔCI) of 90% and 95% confidence intervals. Detailed formulas for these performance indicators are given in Appendix A1.

**Table 1:** Target SI distribution, sample size and censoring scheme considered in the scenarios of the simulation study. The 𝒩(2.8, 2.5^2^) target mimics the SI distribution of SARS-CoV-2 Omicron (Kremer et al., 2022) and the Weibull target 𝒲(2.36, 3.18) mimics the SI distribution of influenza A (Lessler et al., 2009).

| Scenario | SI target distribution | Sample size | Censoring scheme | Results |
| --- | --- | --- | --- | --- |
| 1 | $\mathcal{N}(2.8, 2.5^2)$ | $n = 10$ | Doubly interval-censored | Table 2 |
| 2 | $\mathcal{N}(2.8, 2.5^2)$ | $n = 20$ | Doubly interval-censored | Table 2 |
| 3 | $\mathcal{N}(2.8, 2.5^2)$ | $n = 50$ | Doubly interval-censored | Table 2 |
| 4 | $\mathcal{N}(2.8, 2.5^2)$ | $n = 100$ | Doubly interval-censored | Table 2 |
| 5 | $\mathcal{N}(2.8, 2.5^2)$ | $n = 10$ | Single interval-censored | Table 3 |
| 6 | $\mathcal{N}(2.8, 2.5^2)$ | $n = 20$ | Single interval-censored | Table 3 |
| 7 | $\mathcal{N}(2.8, 2.5^2)$ | $n = 50$ | Single interval-censored | Table 3 |
| 8 | $\mathcal{N}(2.8, 2.5^2)$ | $n = 100$ | Single interval-censored | Table 3 |
| 9 | $\mathcal{N}(2.8, 2.5^2)$ | $n = 10$ | No censoring | Table 4 |
| 10 | $\mathcal{N}(2.8, 2.5^2)$ | $n = 20$ | No censoring | Table 4 |
| 11 | $\mathcal{N}(2.8, 2.5^2)$ | $n = 50$ | No censoring | Table 4 |
| 12 | $\mathcal{N}(2.8, 2.5^2)$ | $n = 100$ | No censoring | Table 4 |
| 13 | $\mathcal{W}(2.36, 3.18)$ | $n = 10$ | Doubly interval-censored | Table 5 |
| 14 | $\mathcal{W}(2.36, 3.18)$ | $n = 20$ | Doubly interval-censored | Table 5 |
| 15 | $\mathcal{W}(2.36, 3.18)$ | $n = 50$ | Doubly interval-censored | Table 5 |
| 16 | $\mathcal{W}(2.36, 3.18)$ | $n = 100$ | Doubly interval-censored | Table 5 |
| 17 | $\mathcal{W}(2.36, 3.18)$ | $n = 10$ | Single interval-censored | Table 6 |
| 18 | $\mathcal{W}(2.36, 3.18)$ | $n = 20$ | Single interval-censored | Table 6 |
| 19 | $\mathcal{W}(2.36, 3.18)$ | $n = 50$ | Single interval-censored | Table 6 |
| 20 | $\mathcal{W}(2.36, 3.18)$ | $n = 100$ | Single interval-censored | Table 6 |
| 21 | $\mathcal{W}(2.36, 3.18)$ | $n = 10$ | No censoring | Table 7 |
| 22 | $\mathcal{W}(2.36, 3.18)$ | $n = 20$ | No censoring | Table 7 |
| 23 | $\mathcal{W}(2.36, 3.18)$ | $n = 50$ | No censoring | Table 7 |
| 24 | $\mathcal{W}(2.36, 3.18)$ | $n = 100$ | No censoring | Table 7 |

**Table 2:** Simulation results for Scenarios 1-4 with a Gaussian target SI distribution *𝒩* (2.8, 2.5^2^) and doubly interval-censored data. The first column contains the selected features of *𝒮*, namely the mean, standard deviation, 5th, 25th, 50th, 75th and 95th percentiles. Bias, ESE, RMSE, coverage probability (CP) and median confidence interval width ΔCI are used to assess the performance of the nonparametric approach.

| **Scenario 1** ($n = 10$) | Bias | ESE | RMSE | CP<sub>90%</sub> | CP<sub>95%</sub> | $\Delta\text{CI}_{90\%}$ | $\Delta\text{CI}_{95\%}$ |
| --- | --- | --- | --- | --- | --- | --- | --- |
| Mean | -0.024 | 0.788 | 0.788 | 86.60 | 93.20 | 2.514 | 2.968 |
| SD | -0.125 | 0.547 | 0.561 | 81.60 | 86.00 | 1.636 | 1.937 |
| $q0.05$ | 0.976 | 1.095 | 1.466 | 59.60 | 64.40 | 2.942 | 3.421 |
| $q0.25$ | 0.160 | 0.875 | 0.889 | 89.20 | 94.60 | 3.372 | 3.970 |
| $q0.50$ | -0.008 | 0.815 | 0.815 | 91.80 | 96.20 | 3.006 | 3.643 |
| $q0.75$ | -0.212 | 0.896 | 0.920 | 89.00 | 92.40 | 3.300 | 3.873 |
| $q0.95$ | -1.040 | 1.116 | 1.524 | 56.20 | 59.40 | 2.935 | 3.463 |
| **Scenario 2** ($n = 20$) | Bias | ESE | RMSE | CP<sub>90%</sub> | CP<sub>95%</sub> | $\Delta\text{CI}_{90\%}$ | $\Delta\text{CI}_{95\%}$ |
| Mean | 0.049 | 0.565 | 0.566 | 88.60 | 93.40 | 1.855 | 2.197 |
| SD | 0.005 | 0.388 | 0.388 | 87.60 | 91.80 | 1.192 | 1.421 |
| $q0.05$ | 0.511 | 0.843 | 0.985 | 76.20 | 80.60 | 2.783 | 3.279 |
| $q0.25$ | 0.081 | 0.653 | 0.658 | 93.00 | 96.40 | 2.439 | 2.929 |
| $q0.50$ | 0.063 | 0.632 | 0.635 | 93.00 | 96.00 | 2.308 | 2.754 |
| $q0.75$ | 0.010 | 0.689 | 0.688 | 89.80 | 95.40 | 2.350 | 2.833 |
| $q0.95$ | -0.430 | 0.842 | 0.945 | 80.80 | 84.60 | 2.785 | 3.251 |
| **Scenario 3** ($n = 50$) | Bias | ESE | RMSE | CP<sub>90%</sub> | CP<sub>95%</sub> | $\Delta\text{CI}_{90\%}$ | $\Delta\text{CI}_{95\%}$ |
| Mean | 0.009 | 0.343 | 0.343 | 92.40 | 95.20 | 1.177 | 1.400 |
| SD | 0.026 | 0.254 | 0.255 | 88.80 | 93.80 | 0.798 | 0.952 |
| $q0.05$ | 0.157 | 0.605 | 0.625 | 88.60 | 93.80 | 2.149 | 2.536 |
| $q0.25$ | 0.024 | 0.424 | 0.424 | 92.60 | 96.80 | 1.556 | 1.847 |
| $q0.50$ | 0.004 | 0.374 | 0.374 | 94.20 | 98.00 | 1.428 | 1.710 |
| $q0.75$ | -0.008 | 0.421 | 0.421 | 93.60 | 96.60 | 1.563 | 1.864 |
| $q0.95$ | -0.124 | 0.613 | 0.625 | 86.60 | 92.40 | 2.106 | 2.527 |
| **Scenario 4** ($n = 100$) | Bias | ESE | RMSE | CP<sub>90%</sub> | CP<sub>95%</sub> | $\Delta\text{CI}_{90\%}$ | $\Delta\text{CI}_{95\%}$ |
| Mean | 0.002 | 0.254 | 0.253 | 89.00 | 95.40 | 0.841 | 1.006 |
| SD | 0.061 | 0.175 | 0.185 | 90.20 | 94.80 | 0.586 | 0.700 |
| $q0.05$ | -0.007 | 0.444 | 0.443 | 92.60 | 96.60 | 1.634 | 1.971 |
| $q0.25$ | -0.021 | 0.313 | 0.313 | 92.20 | 96.80 | 1.136 | 1.355 |
| $q0.50$ | 0.010 | 0.287 | 0.287 | 94.00 | 97.60 | 1.038 | 1.240 |
| $q0.75$ | 0.028 | 0.309 | 0.310 | 92.20 | 96.60 | 1.118 | 1.335 |
| $q0.95$ | -0.001 | 0.435 | 0.435 | 92.20 | 96.40 | 1.665 | 1.986 |

**Table 3:** Simulation results for Scenarios 5-8 with a Gaussian target SI distribution *𝒩* (2.8, 2.5^2^) and single interval-censored data. The first column contains the selected features of *S*, namely the mean, standard deviation, 5th, 25th, 50th, 75th and 95th percentiles. Bias, ESE, RMSE, coverage probability (CP) and median confidence interval width ΔCI are used to assess the performance of the nonparametric approach.

| **Scenario 5** ($n = 10$) | Bias | ESE | RMSE | CP<sub>90%</sub> | CP<sub>95%</sub> | $\Delta\text{CI}_{90\%}$ | $\Delta\text{CI}_{95\%}$ |
| --- | --- | --- | --- | --- | --- | --- | --- |
| Mean | 0.006 | 0.847 | 0.847 | 83.00 | 88.40 | 2.422 | 2.866 |
| SD | -0.226 | 0.512 | 0.559 | 77.40 | 82.60 | 1.546 | 1.818 |
| $q0.05$ | 1.168 | 1.104 | 1.607 | 50.00 | 51.60 | 2.716 | 3.113 |
| $q0.25$ | 0.272 | 0.934 | 0.972 | 88.60 | 93.00 | 3.308 | 3.785 |
| $q0.50$ | 0.011 | 0.910 | 0.909 | 83.40 | 92.00 | 2.908 | 3.599 |
| $q0.75$ | -0.250 | 0.942 | 0.974 | 86.40 | 89.60 | 3.232 | 3.701 |
| $q0.95$ | -1.174 | 1.126 | 1.626 | 45.20 | 49.00 | 2.619 | 3.004 |
| **Scenario 6** ($n = 20$) | Bias | ESE | RMSE | CP<sub>90%</sub> | CP<sub>95%</sub> | $\Delta\text{CI}_{90\%}$ | $\Delta\text{CI}_{95\%}$ |
| Mean | -0.037 | 0.575 | 0.575 | 87.20 | 92.80 | 1.778 | 2.115 |
| SD | -0.090 | 0.409 | 0.419 | 81.20 | 87.20 | 1.177 | 1.394 |
| $q0.05$ | 0.552 | 0.904 | 1.059 | 71.20 | 74.00 | 2.619 | 2.979 |
| $q0.25$ | 0.076 | 0.691 | 0.695 | 88.00 | 93.00 | 2.344 | 2.796 |
| $q0.50$ | -0.015 | 0.618 | 0.618 | 90.40 | 95.20 | 2.155 | 2.600 |
| $q0.75$ | -0.156 | 0.652 | 0.669 | 87.80 | 92.60 | 2.271 | 2.758 |
| $q0.95$ | -0.651 | 0.913 | 1.120 | 68.80 | 72.40 | 2.670 | 3.030 |
| **Scenario 7** ($n = 50$) | Bias | ESE | RMSE | CP<sub>90%</sub> | CP<sub>95%</sub> | $\Delta\text{CI}_{90\%}$ | $\Delta\text{CI}_{95\%}$ |
| Mean | 0.004 | 0.354 | 0.353 | 88.40 | 94.20 | 1.148 | 1.370 |
| SD | -0.027 | 0.254 | 0.255 | 86.80 | 92.00 | 0.783 | 0.932 |
| $q0.05$ | 0.220 | 0.632 | 0.669 | 84.40 | 87.60 | 2.168 | 2.575 |
| $q0.25$ | 0.053 | 0.429 | 0.432 | 90.80 | 95.80 | 1.520 | 1.815 |
| $q0.50$ | 0.013 | 0.403 | 0.403 | 90.00 | 95.40 | 1.405 | 1.696 |
| $q0.75$ | -0.033 | 0.440 | 0.441 | 89.40 | 94.40 | 1.514 | 1.821 |
| $q0.95$ | -0.238 | 0.619 | 0.662 | 83.60 | 88.40 | 2.160 | 2.510 |
| **Scenario 8** ($n = 100$) | Bias | ESE | RMSE | CP<sub>90%</sub> | CP<sub>95%</sub> | $\Delta\text{CI}_{90\%}$ | $\Delta\text{CI}_{95\%}$ |
| Mean | 0.010 | 0.248 | 0.248 | 88.80 | 94.40 | 0.826 | 0.985 |
| SD | 0.007 | 0.173 | 0.173 | 89.20 | 93.80 | 0.563 | 0.671 |
| $q0.05$ | 0.094 | 0.447 | 0.457 | 88.80 | 93.00 | 1.585 | 1.889 |
| $q0.25$ | 0.016 | 0.320 | 0.320 | 92.20 | 95.80 | 1.113 | 1.336 |
| $q0.50$ | 0.005 | 0.300 | 0.300 | 89.60 | 96.20 | 1.012 | 1.212 |
| $q0.75$ | 0.011 | 0.310 | 0.310 | 92.00 | 97.00 | 1.108 | 1.322 |
| $q0.95$ | -0.076 | 0.418 | 0.425 | 88.60 | 93.60 | 1.568 | 1.871 |

**Table 4:** Simulation results for Scenarios 9-12 with a Gaussian target SI distribution *𝒩* (2.8, 2.5^2^) and exactly observed data. The first column contains the selected features of *𝒮*, namely the mean, standard deviation, 5th, 25th, 50th, 75th and 95th percentiles. Bias, ESE, RMSE, coverage probability (CP) and median confidence interval width ΔCI are used to assess the performance of the nonparametric approach.

| **Scenario 9** ($n = 10$) | Bias | ESE | RMSE | CP<sub>90%</sub> | CP<sub>95%</sub> | $\Delta\text{CI}_{90\%}$ | $\Delta\text{CI}_{95\%}$ |
| --- | --- | --- | --- | --- | --- | --- | --- |
| Mean | -0.024 | 0.781 | 0.781 | 87.20 | 89.20 | 2.402 | 2.850 |
| SD | -0.196 | 0.543 | 0.576 | 78.00 | 83.20 | 1.589 | 1.887 |
| $q0.05$ | 1.092 | 1.113 | 1.558 | 51.60 | 53.80 | 2.772 | 3.190 |
| $q0.25$ | 0.216 | 0.885 | 0.910 | 89.40 | 92.20 | 3.455 | 3.901 |
| $q0.50$ | -0.007 | 0.830 | 0.829 | 89.40 | 93.80 | 2.925 | 3.606 |
| $q0.75$ | -0.249 | 0.891 | 0.924 | 88.80 | 91.60 | 3.297 | 3.782 |
| $q0.95$ | -1.176 | 1.103 | 1.612 | 46.60 | 49.00 | 2.582 | 3.030 |
| **Scenario 10** ($n = 20$) | Bias | ESE | RMSE | CP<sub>90%</sub> | CP<sub>95%</sub> | $\Delta\text{CI}_{90\%}$ | $\Delta\text{CI}_{95\%}$ |
| Mean | -0.010 | 0.577 | 0.576 | 87.80 | 93.60 | 1.760 | 2.088 |
| SD | -0.112 | 0.369 | 0.385 | 82.00 | 87.60 | 1.159 | 1.378 |
| $q0.05$ | 0.631 | 0.880 | 1.082 | 67.80 | 70.20 | 2.627 | 3.013 |
| $q0.25$ | 0.118 | 0.702 | 0.711 | 86.60 | 92.40 | 2.306 | 2.709 |
| $q0.50$ | -0.011 | 0.637 | 0.637 | 89.60 | 94.20 | 2.145 | 2.554 |
| $q0.75$ | -0.147 | 0.639 | 0.656 | 90.00 | 93.00 | 2.266 | 2.730 |
| $q0.95$ | -0.641 | 0.847 | 1.061 | 69.80 | 72.60 | 2.702 | 3.042 |
| **Scenario 11** ($n = 50$) | Bias | ESE | RMSE | CP<sub>90%</sub> | CP<sub>95%</sub> | $\Delta\text{CI}_{90\%}$ | $\Delta\text{CI}_{95\%}$ |
| Mean | -0.007 | 0.350 | 0.350 | 89.20 | 94.60 | 1.159 | 1.380 |
| SD | -0.020 | 0.247 | 0.247 | 87.20 | 93.00 | 0.795 | 0.943 |
| $q0.05$ | 0.233 | 0.615 | 0.657 | 85.60 | 90.20 | 2.186 | 2.532 |
| $q0.25$ | 0.036 | 0.431 | 0.432 | 90.40 | 94.40 | 1.500 | 1.791 |
| $q0.50$ | -0.032 | 0.403 | 0.404 | 90.20 | 94.60 | 1.400 | 1.672 |
| $q0.75$ | -0.060 | 0.447 | 0.451 | 89.60 | 95.60 | 1.545 | 1.858 |
| $q0.95$ | -0.184 | 0.600 | 0.627 | 88.40 | 91.20 | 2.198 | 2.564 |
| **Scenario 12** ($n = 100$) | Bias | ESE | RMSE | CP<sub>90%</sub> | CP<sub>95%</sub> | $\Delta\text{CI}_{90\%}$ | $\Delta\text{CI}_{95\%}$ |
| Mean | 0.011 | 0.257 | 0.257 | 88.20 | 94.00 | 0.825 | 0.981 |
| SD | 0.009 | 0.171 | 0.171 | 91.60 | 95.60 | 0.571 | 0.679 |
| $q0.05$ | 0.097 | 0.441 | 0.451 | 88.80 | 93.40 | 1.593 | 1.909 |
| $q0.25$ | 0.013 | 0.323 | 0.323 | 90.80 | 96.00 | 1.109 | 1.332 |
| $q0.50$ | 0.024 | 0.297 | 0.298 | 91.40 | 95.60 | 1.017 | 1.218 |
| $q0.75$ | 0.002 | 0.322 | 0.322 | 89.40 | 95.80 | 1.076 | 1.287 |
| $q0.95$ | -0.083 | 0.444 | 0.451 | 90.00 | 94.00 | 1.620 | 1.924 |

**Table 5:** Simulation results for Scenarios 13-16 with a Weibull target SI distribution 𝒲 (2.36, 3.18) and doubly interval-censored data. The first column contains the selected features of *𝒮*, namely the mean, standard deviation, 5th, 25th, 50th, 75th and 95th percentiles. Bias, ESE, RMSE, coverage probability (CP) and median confidence interval width ΔCI are used to assess the performance of the nonparametric approach.

| **Scenario 13** ($n = 10$) | Bias | ESE | RMSE | CP<sub>90%</sub> | CP<sub>95%</sub> | $\Delta\text{CI}_{90\%}$ | $\Delta\text{CI}_{95\%}$ |
| --- | --- | --- | --- | --- | --- | --- | --- |
| Mean | -0.007 | 0.425 | 0.425 | 86.60 | 93.20 | 1.378 | 1.648 |
| SD | 0.049 | 0.250 | 0.254 | 94.40 | 97.40 | 0.930 | 1.099 |
| $q0.05$ | 0.232 | 0.426 | 0.485 | 85.00 | 89.40 | 1.613 | 1.900 |
| $q0.25$ | 0.088 | 0.427 | 0.436 | 93.60 | 97.60 | 1.642 | 1.958 |
| $q0.50$ | 0.033 | 0.448 | 0.448 | 91.80 | 96.00 | 1.667 | 2.002 |
| $q0.75$ | -0.041 | 0.504 | 0.505 | 90.80 | 96.20 | 1.895 | 2.226 |
| $q0.95$ | -0.453 | 0.640 | 0.784 | 68.60 | 74.00 | 1.947 | 2.267 |
| **Scenario 14** ($n = 20$) | Bias | ESE | RMSE | CP<sub>90%</sub> | CP<sub>95%</sub> | $\Delta\text{CI}_{90\%}$ | $\Delta\text{CI}_{95\%}$ |
| Mean | -0.014 | 0.286 | 0.286 | 91.40 | 94.60 | 1.024 | 1.217 |
| SD | 0.112 | 0.178 | 0.210 | 94.20 | 97.80 | 0.673 | 0.797 |
| $q0.05$ | -0.010 | 0.298 | 0.298 | 96.60 | 99.20 | 1.422 | 1.690 |
| $q0.25$ | -0.014 | 0.294 | 0.294 | 96.00 | 98.20 | 1.244 | 1.488 |
| $q0.50$ | 0.017 | 0.313 | 0.313 | 95.00 | 98.60 | 1.280 | 1.528 |
| $q0.75$ | 0.032 | 0.361 | 0.362 | 94.40 | 97.60 | 1.422 | 1.696 |
| $q0.95$ | -0.166 | 0.473 | 0.501 | 87.60 | 92.20 | 1.786 | 2.123 |
| **Scenario 15** ($n = 50$) | Bias | ESE | RMSE | CP<sub>90%</sub> | CP<sub>95%</sub> | $\Delta\text{CI}_{90\%}$ | $\Delta\text{CI}_{95\%}$ |
| Mean | -0.001 | 0.184 | 0.184 | 92.40 | 95.80 | 0.657 | 0.784 |
| SD | 0.136 | 0.115 | 0.178 | 83.80 | 92.80 | 0.443 | 0.528 |
| $q0.05$ | -0.165 | 0.204 | 0.262 | 93.60 | 98.40 | 0.975 | 1.160 |
| $q0.25$ | -0.044 | 0.195 | 0.200 | 95.60 | 98.40 | 0.822 | 0.975 |
| $q0.50$ | 0.024 | 0.203 | 0.205 | 94.80 | 97.40 | 0.833 | 0.992 |
| $q0.75$ | 0.073 | 0.229 | 0.240 | 94.80 | 98.60 | 0.934 | 1.120 |
| $q0.95$ | 0.067 | 0.332 | 0.339 | 94.60 | 96.60 | 1.327 | 1.581 |
| **Scenario 16** ($n = 100$) | Bias | ESE | RMSE | CP<sub>90%</sub> | CP<sub>95%</sub> | $\Delta\text{CI}_{90\%}$ | $\Delta\text{CI}_{95\%}$ |
| Mean | -0.001 | 0.128 | 0.128 | 94.00 | 97.40 | 0.470 | 0.561 |
| SD | 0.154 | 0.081 | 0.174 | 54.40 | 68.40 | 0.323 | 0.385 |
| $q0.05$ | -0.234 | 0.145 | 0.275 | 84.60 | 93.20 | 0.737 | 0.876 |
| $q0.25$ | -0.065 | 0.137 | 0.152 | 94.60 | 97.80 | 0.586 | 0.701 |
| $q0.50$ | 0.017 | 0.143 | 0.144 | 97.20 | 98.80 | 0.591 | 0.706 |
| $q0.75$ | 0.091 | 0.162 | 0.186 | 93.80 | 97.20 | 0.684 | 0.817 |
| $q0.95$ | 0.160 | 0.238 | 0.287 | 95.20 | 98.60 | 1.048 | 1.241 |

**Table 6:** Simulation results for Scenarios 17-20 with a Weibull target SI distribution *𝒲* (2.36, 3.18) and single interval-censored data. The first column contains the selected features of *𝒮*, namely the mean, standard deviation, 5th, 25th, 50th, 75th and 95th percentiles. Bias, ESE, RMSE, coverage probability (CP) and median confidence interval width ΔCI are used to assess the performance of the nonparametric approach.

| **Scenario 17** ($n = 10$) | Bias | ESE | RMSE | CP<sub>90%</sub> | CP<sub>95%</sub> | $\Delta\text{CI}_{90\%}$ | $\Delta\text{CI}_{95\%}$ |
| --- | --- | --- | --- | --- | --- | --- | --- |
| Mean | -0.005 | 0.415 | 0.415 | 87.00 | 93.00 | 1.289 | 1.533 |
| SD | -0.050 | 0.279 | 0.283 | 83.60 | 87.60 | 0.824 | 0.973 |
| $q0.05$ | 0.402 | 0.411 | 0.575 | 66.20 | 70.00 | 1.283 | 1.499 |
| $q0.25$ | 0.132 | 0.412 | 0.432 | 89.00 | 93.20 | 1.502 | 1.792 |
| $q0.50$ | 0.022 | 0.452 | 0.452 | 89.80 | 94.60 | 1.572 | 1.902 |
| $q0.75$ | -0.099 | 0.530 | 0.539 | 88.60 | 92.60 | 1.873 | 2.169 |
| $q0.95$ | -0.583 | 0.668 | 0.886 | 55.80 | 59.60 | 1.669 | 1.946 |
| **Scenario 18** ($n = 20$) | Bias | ESE | RMSE | CP<sub>90%</sub> | CP<sub>95%</sub> | $\Delta\text{CI}_{90\%}$ | $\Delta\text{CI}_{95\%}$ |
| Mean | 0.007 | 0.281 | 0.281 | 88.60 | 93.40 | 0.921 | 1.099 |
| SD | -0.016 | 0.202 | 0.202 | 84.60 | 90.20 | 0.602 | 0.715 |
| $q0.05$ | 0.205 | 0.322 | 0.381 | 82.40 | 86.40 | 1.166 | 1.387 |
| $q0.25$ | 0.085 | 0.316 | 0.327 | 89.40 | 94.60 | 1.146 | 1.359 |
| $q0.50$ | 0.035 | 0.327 | 0.328 | 90.20 | 95.20 | 1.135 | 1.363 |
| $q0.75$ | -0.036 | 0.359 | 0.361 | 90.80 | 94.60 | 1.303 | 1.554 |
| $q0.95$ | -0.316 | 0.499 | 0.591 | 73.80 | 77.60 | 1.604 | 1.851 |
| **Scenario 19** ($n = 50$) | Bias | ESE | RMSE | CP<sub>90%</sub> | CP<sub>95%</sub> | $\Delta\text{CI}_{90\%}$ | $\Delta\text{CI}_{95\%}$ |
| Mean | -0.012 | 0.166 | 0.166 | 92.40 | 97.20 | 0.608 | 0.723 |
| SD | 0.029 | 0.118 | 0.121 | 91.60 | 95.20 | 0.401 | 0.476 |
| $q0.05$ | 0.010 | 0.190 | 0.190 | 95.80 | 98.00 | 0.794 | 0.947 |
| $q0.25$ | -0.005 | 0.176 | 0.176 | 95.40 | 97.60 | 0.743 | 0.887 |
| $q0.50$ | 0.007 | 0.193 | 0.193 | 95.00 | 98.80 | 0.769 | 0.924 |
| $q0.75$ | 0.002 | 0.232 | 0.231 | 94.80 | 97.60 | 0.873 | 1.046 |
| $q0.95$ | -0.102 | 0.347 | 0.361 | 86.40 | 91.00 | 1.254 | 1.511 |
| **Scenario 20** ($n = 100$) | Bias | ESE | RMSE | CP<sub>90%</sub> | CP<sub>95%</sub> | $\Delta\text{CI}_{90\%}$ | $\Delta\text{CI}_{95\%}$ |
| Mean | 0.002 | 0.127 | 0.127 | 91.00 | 95.40 | 0.432 | 0.514 |
| SD | 0.040 | 0.088 | 0.097 | 89.00 | 94.20 | 0.291 | 0.345 |
| $q0.05$ | -0.023 | 0.146 | 0.147 | 95.20 | 98.60 | 0.603 | 0.715 |
| $q0.25$ | -0.006 | 0.143 | 0.143 | 92.80 | 95.80 | 0.533 | 0.639 |
| $q0.50$ | 0.011 | 0.152 | 0.152 | 93.40 | 96.80 | 0.548 | 0.657 |
| $q0.75$ | 0.019 | 0.176 | 0.177 | 91.60 | 96.00 | 0.630 | 0.756 |
| $q0.95$ | 0.002 | 0.262 | 0.262 | 91.80 | 96.00 | 0.976 | 1.163 |

**Table 7:**
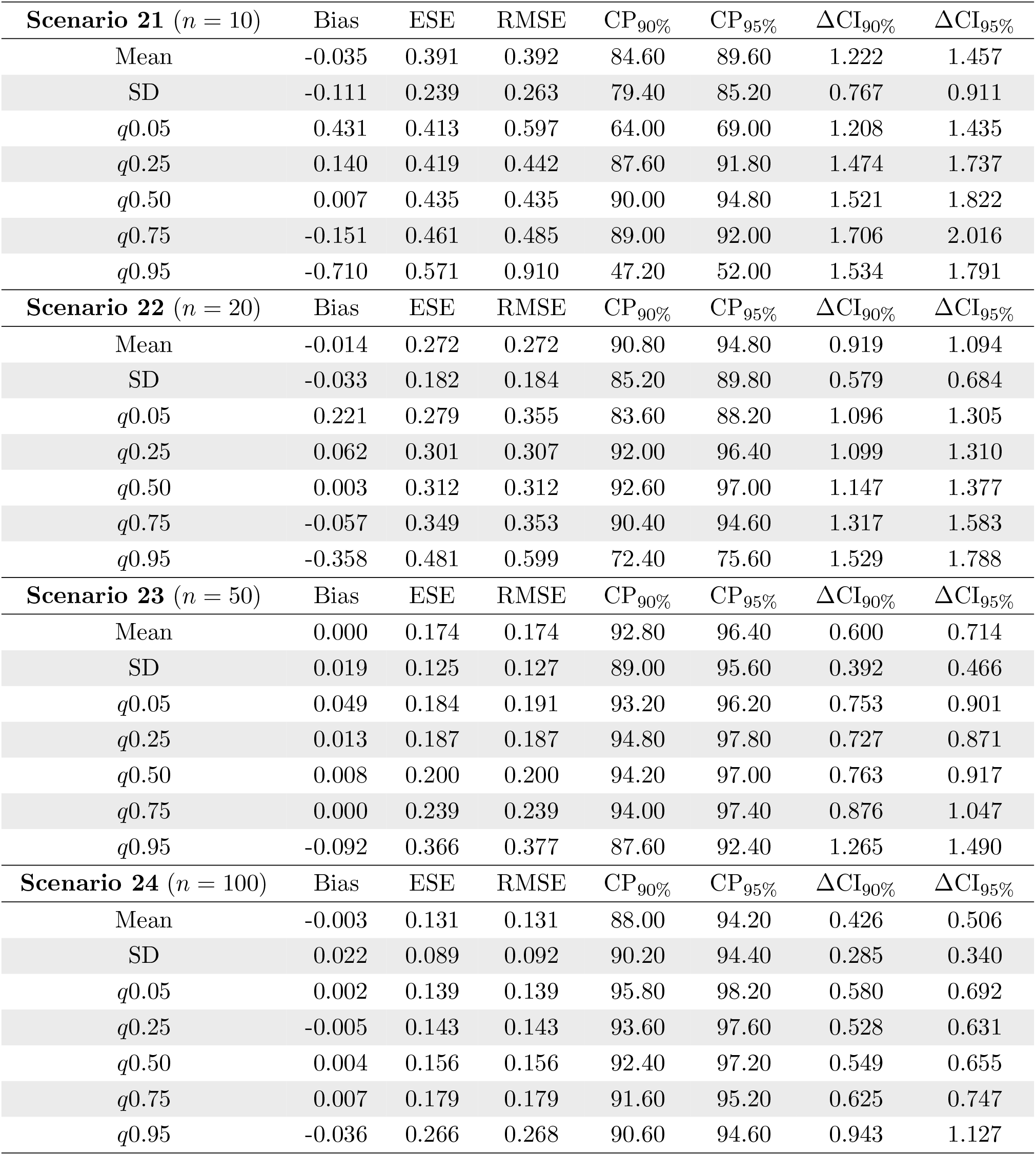
Simulation results for Scenarios 21-24 with a Weibull target SI distribution *𝒲* (2.36, 3.18) and exactly observed data. The first column contains the selected features of *𝒮*, namely the mean, standard deviation, 5th, 25th, 50th, 75th and 95th percentiles. Bias, ESE, RMSE, coverage probability (CP) and median confidence interval width ΔCI are used to assess the performance of the nonparametric approach.

| **Scenario 21** ($n = 10$) | Bias | ESE | RMSE | CP<sub>90%</sub> | CP<sub>95%</sub> | $\Delta\text{CI}_{90\%}$ | $\Delta\text{CI}_{95\%}$ |
| --- | --- | --- | --- | --- | --- | --- | --- |
| Mean | -0.035 | 0.391 | 0.392 | 84.60 | 89.60 | 1.222 | 1.457 |
| SD | -0.111 | 0.239 | 0.263 | 79.40 | 85.20 | 0.767 | 0.911 |
| $q0.05$ | 0.431 | 0.413 | 0.597 | 64.00 | 69.00 | 1.208 | 1.435 |
| $q0.25$ | 0.140 | 0.419 | 0.442 | 87.60 | 91.80 | 1.474 | 1.737 |
| $q0.50$ | 0.007 | 0.435 | 0.435 | 90.00 | 94.80 | 1.521 | 1.822 |
| $q0.75$ | -0.151 | 0.461 | 0.485 | 89.00 | 92.00 | 1.706 | 2.016 |
| $q0.95$ | -0.710 | 0.571 | 0.910 | 47.20 | 52.00 | 1.534 | 1.791 |
| **Scenario 22** ($n = 20$) | Bias | ESE | RMSE | CP<sub>90%</sub> | CP<sub>95%</sub> | $\Delta\text{CI}_{90\%}$ | $\Delta\text{CI}_{95\%}$ |
| Mean | -0.014 | 0.272 | 0.272 | 90.80 | 94.80 | 0.919 | 1.094 |
| SD | -0.033 | 0.182 | 0.184 | 85.20 | 89.80 | 0.579 | 0.684 |
| $q0.05$ | 0.221 | 0.279 | 0.355 | 83.60 | 88.20 | 1.096 | 1.305 |
| $q0.25$ | 0.062 | 0.301 | 0.307 | 92.00 | 96.40 | 1.099 | 1.310 |
| $q0.50$ | 0.003 | 0.312 | 0.312 | 92.60 | 97.00 | 1.147 | 1.377 |
| $q0.75$ | -0.057 | 0.349 | 0.353 | 90.40 | 94.60 | 1.317 | 1.583 |
| $q0.95$ | -0.358 | 0.481 | 0.599 | 72.40 | 75.60 | 1.529 | 1.788 |
| **Scenario 23** ($n = 50$) | Bias | ESE | RMSE | CP<sub>90%</sub> | CP<sub>95%</sub> | $\Delta\text{CI}_{90\%}$ | $\Delta\text{CI}_{95\%}$ |
| Mean | 0.000 | 0.174 | 0.174 | 92.80 | 96.40 | 0.600 | 0.714 |
| SD | 0.019 | 0.125 | 0.127 | 89.00 | 95.60 | 0.392 | 0.466 |
| $q0.05$ | 0.049 | 0.184 | 0.191 | 93.20 | 96.20 | 0.753 | 0.901 |
| $q0.25$ | 0.013 | 0.187 | 0.187 | 94.80 | 97.80 | 0.727 | 0.871 |
| $q0.50$ | 0.008 | 0.200 | 0.200 | 94.20 | 97.00 | 0.763 | 0.917 |
| $q0.75$ | 0.000 | 0.239 | 0.239 | 94.00 | 97.40 | 0.876 | 1.047 |
| $q0.95$ | -0.092 | 0.366 | 0.377 | 87.60 | 92.40 | 1.265 | 1.490 |
| **Scenario 24** ($n = 100$) | Bias | ESE | RMSE | CP<sub>90%</sub> | CP<sub>95%</sub> | $\Delta\text{CI}_{90\%}$ | $\Delta\text{CI}_{95\%}$ |
| Mean | -0.003 | 0.131 | 0.131 | 88.00 | 94.20 | 0.426 | 0.506 |
| SD | 0.022 | 0.089 | 0.092 | 90.20 | 94.40 | 0.285 | 0.340 |
| $q0.05$ | 0.002 | 0.139 | 0.139 | 95.80 | 98.20 | 0.580 | 0.692 |
| $q0.25$ | -0.005 | 0.143 | 0.143 | 93.60 | 97.60 | 0.528 | 0.631 |
| $q0.50$ | 0.004 | 0.156 | 0.156 | 92.40 | 97.20 | 0.549 | 0.655 |
| $q0.75$ | 0.007 | 0.179 | 0.179 | 91.60 | 95.20 | 0.625 | 0.747 |
| $q0.95$ | -0.036 | 0.266 | 0.268 | 90.60 | 94.60 | 0.943 | 1.127 |

Scenarios imitating SARS-CoV-2 serial interval data (Scenarios 1-12) show that the bias has a tendency to decrease with increasing sample size. Without surprise, the bias is largest for the 5th and 95th percentiles for small to medium sample size (*n* ≤ 50) since information carried by the data is not rich enough to accurately capture the tail behavior of the target SI distribution. For large sample size (*n* = 100), the bias becomes negligible, even for percentiles in the tails. Moreover, the ESE and RMSE systematically decline as the sample size increases. Coverage probability of the 90% and 95% confidence interval, respectively, tends to come closer to its respective nominal value as the sample size increases.

Note that even under small to moderate sample size, the coverage probability is reasonably close to its nominal level, except for the 5th and 95th percentiles, where undercoverage is observed. Globally, the median width of confidence intervals obtained with the percentile bootstrap method tends to decrease with increasing sample size.

Similar interpretations of the simulation results can be made for the scenarios mimicking influenza A serial interval data (Scenarios 13-24). For doubly interval-censored data, our method has difficulties to estimate the standard deviation (Scenarios 13-16) and confidence intervals tend to undercover. This phenomenon vanishes when considering single interval-censored data or exactly observed serial interval data.

## 4 Applications to real serial interval data

We illustrate our nonparametric approach on five real serial interval datasets that are publicly available. Results can be reproduced with code available on the GitHub repository based on the EpiLPS package (https://github.com/oswaldogressani/Serial_interval).

### 4.1 Influenza A (2009 H1N1 influenza) at a New York City school

We start by analyzing a dataset based on illness onset dates of *n* = 16 infector-infectee pairs obtained from the supplementary appendix of Lessler et al. (2009). After fitting a Weibull distribution to the data, the authors obtain a median serial interval of 2.7 days (CI95% 2.0-3.5) and a 95th percentile of 5.1 days (CI95% 3.6-6.5). Our nonparametric method estimates that the median SI is 2.8 days (CI95% 1.6-4.0) and the 95th percentile estimate is 4.9 days (CI95% 4.1-5.8). Figure 1 summarizes the observed serial interval windows and the point and interval estimates of selected features of the serial interval *S*. The light blue curves represent smoothed estimates of the cdf of *S* for *B* = 5000 bootstrap samples, where smoothing is implemented with the Laplacian-P-splines methodology (Gressani and Lambert, 2018, 2021).

**Figure 1.**
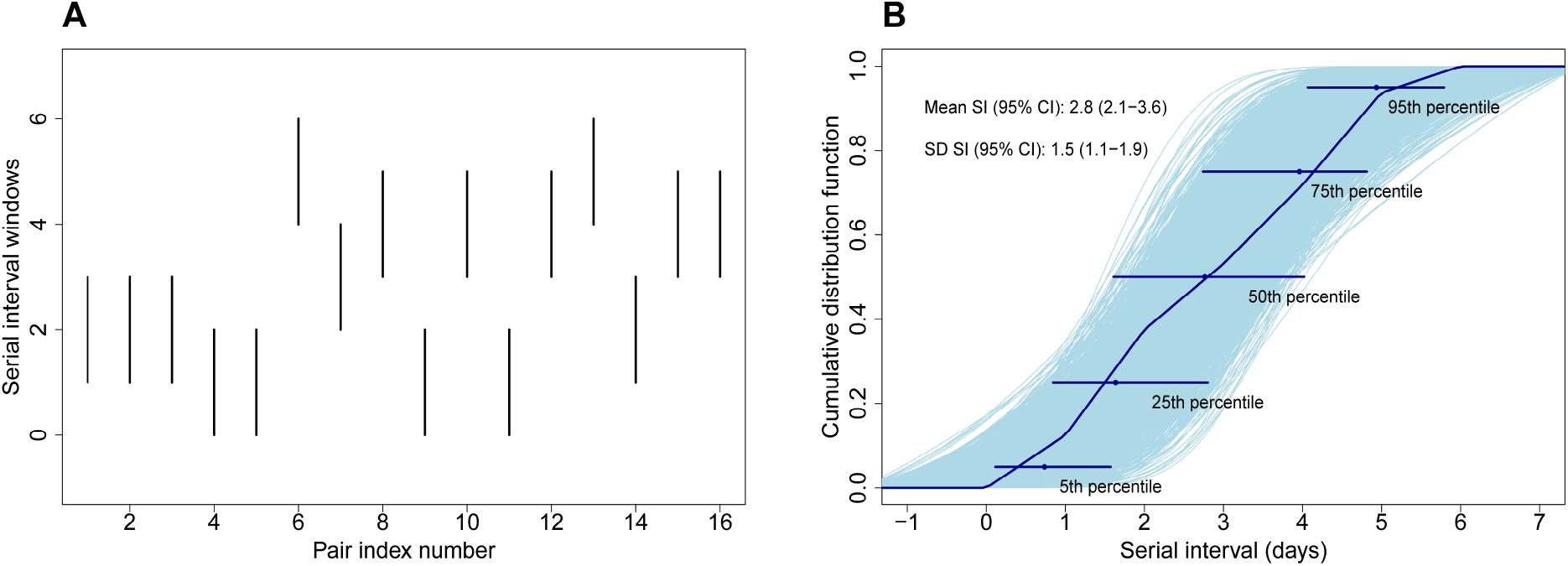
(**A**) Serial interval windows of influenza A for *n* = 16 infector-infectee pairs at a New York City school (Lessler et al., 2009). (**B**) Nonparametric estimate 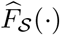 (dark blue); smoothed estimates of the cdf of *S* for *B* = 5000 bootstrap samples (light blue) and 95% CIs for selected percentiles with associated point estimate (dark blue dot).

### 4.2 Influenza A (2009 H1N1 influenza) in San Antonio, Texas, USA

Another dataset on influenza is downloaded from the EpiEstim package (Cori et al., 2013) and contains doubly interval-censored serial interval data from the 2009 influenza A outbreak in San Antonio, Texas, USA (Morgan et al., 2010). Based on our nonparametric methodology, EpiLPS estimates a mean serial interval of 4.0 days (CI95% 3.1-5.0). The standard deviation of the serial interval is estimated at 1.9 days (CI95% 1.2-2.6) and the 95th percentile is at 7.0 days (CI95% 5.0-8.7). Serial interval windows and estimates of different features of *S* are shown in Figure 2.

**Figure 2.**
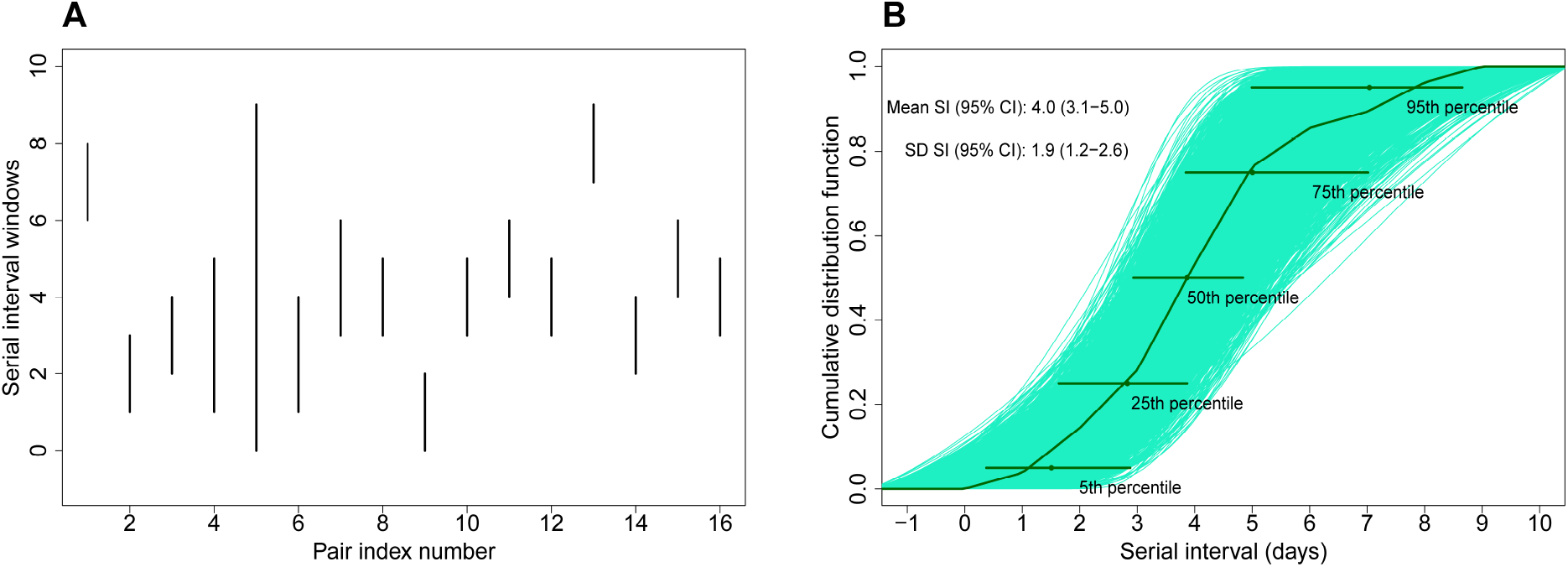
(**A**) Serial interval windows of influenza A for *n* = 16 infector-infectee pairs in San Antonio, Texas, USA (Cori et al., 2013). (**B**) Nonparametric estimate 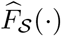 (dark green); smoothed estimates of the cdf of *𝒮* for *B* = 5000 bootstrap samples (light green) and 95% CIs for selected percentiles with associated point estimate (dark green dot).

### 4.3 Illness onset data for 2019-nCoV in Wuhan, China

Li et al. (2020) share data on illness onset dates of *n* = 6 infector-infectee pairs and estimate that the serial interval has a mean of 7.5 days (CI95% 5.3-19) based on a parametric model involving a Gamma distribution. EpiLPS obtains a mean serial interval estimate of 6.3 days (CI95% 4.7-7.8) and a median SI of 6.5 days (CI95% 4.1-8.0).

### 4.4 Illness onset data for 2019-nCoV with *n* = 28 infector-infectee pairs

A richer serial interval dataset on 2019-nCoV is provided by Nishiura et al. (2020). They obtained doubly interval-censored data on *n* = 28 infector-infectee pairs and estimated features of the serial interval based on a Bayesian parametric approach. The authors estimate the median serial interval to be 4.0 days (CrI95% 3.1-4.9), where CrI denotes the credible interval. The mean and standard deviation of the serial interval are estimated at 4.7 days (CrI95% 3.7-6.0) and 2.9 days (CrI95% 1.9-4.9), respectively. Our nonparametric method estimates the median serial interval at 3.8 days (CI95% 3.1-4.9). Also, EpiLPS estimates the mean and standard deviation of the serial interval at 4.6 days (CI95% 3.7-5.6) and 2.6 days (CI95% 1.9-3.2), respectively. A graphical output of the EpiLPS results is shown in Figure 3.

**Figure 3.**
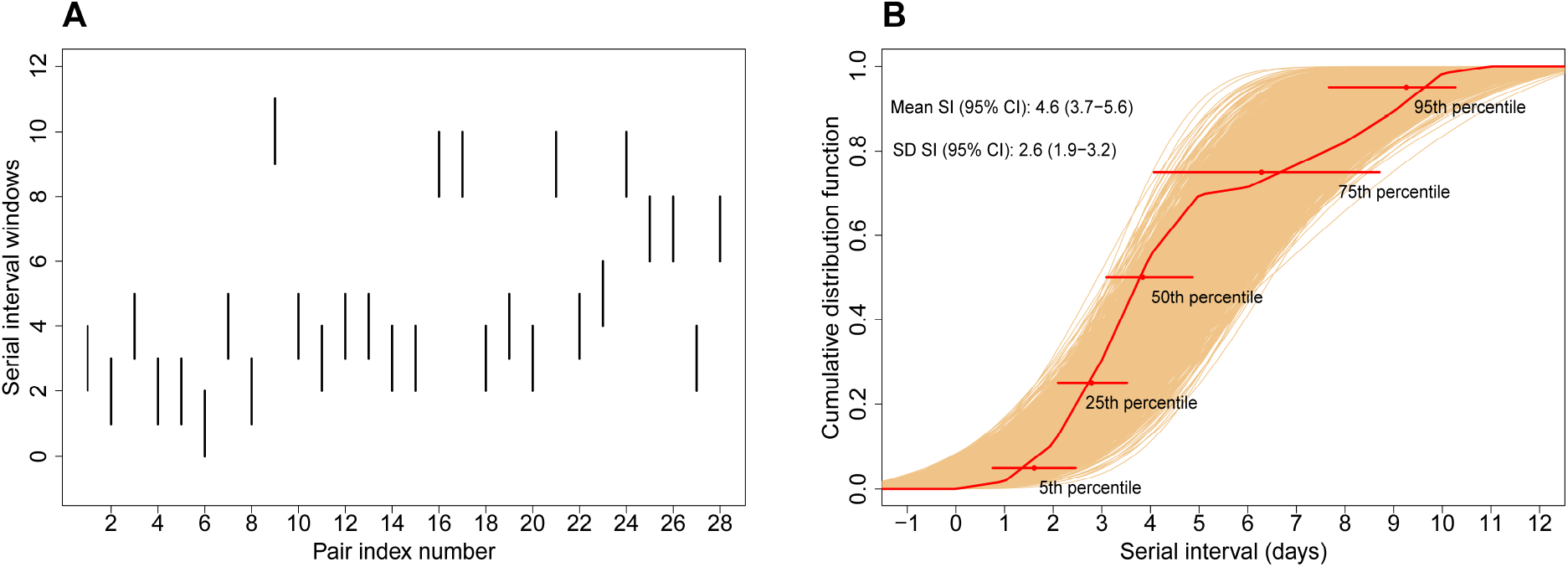
(**A**) Serial interval windows of 2019-nCoV for *n* = 28 infector-infectee pairs (Nishiura et al., 2020). (**B**) Nonparametric estimate 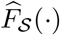(red); smoothed estimates of the cdf of *S* for *B* = 5000 bootstrap samples (orange) and 95% CIs for selected percentiles with associated point estimate (red dot).

### 4.5 Illness onset data for SARS-CoV-2 in Belgium

Kremer et al. (2022) report data on illness onset dates of *n* = 2161 transmission pairs for the Omicron variant of SARS-CoV-2 and *n* = 334 infector-infectee pairs for the Delta variant. Fitting a Gaussian distribution to the data using a Bayesian approach, the authors obtain a median serial interval of 2.75 days (CrI95% 2.65-2.86) and a standard deviation of 2.54 days (CrI95% 2.46-2.61) for Omicron. For Delta, they obtain a median serial interval of 3.00 days (CrI95% 2.73-3.26) and a standard deviation of 2.49 days (CrI95% 2.31-2.69). With our nonparametric approach in EpiLPS, we obtain an estimated median SI at 2.62 days (CI95% 2.50-2.74) and a standard deviation of 2.55 days (CI95% 2.46-2.64) for Omicron. For Delta, EpiLPS estimates the median SI at 3.05 days (CI95% 2.76-3.34) and the estimated standard deviation is 2.49 days (CI95% 2.30-2.69). Results are summarized in Figure 4.

**Figure 4.**
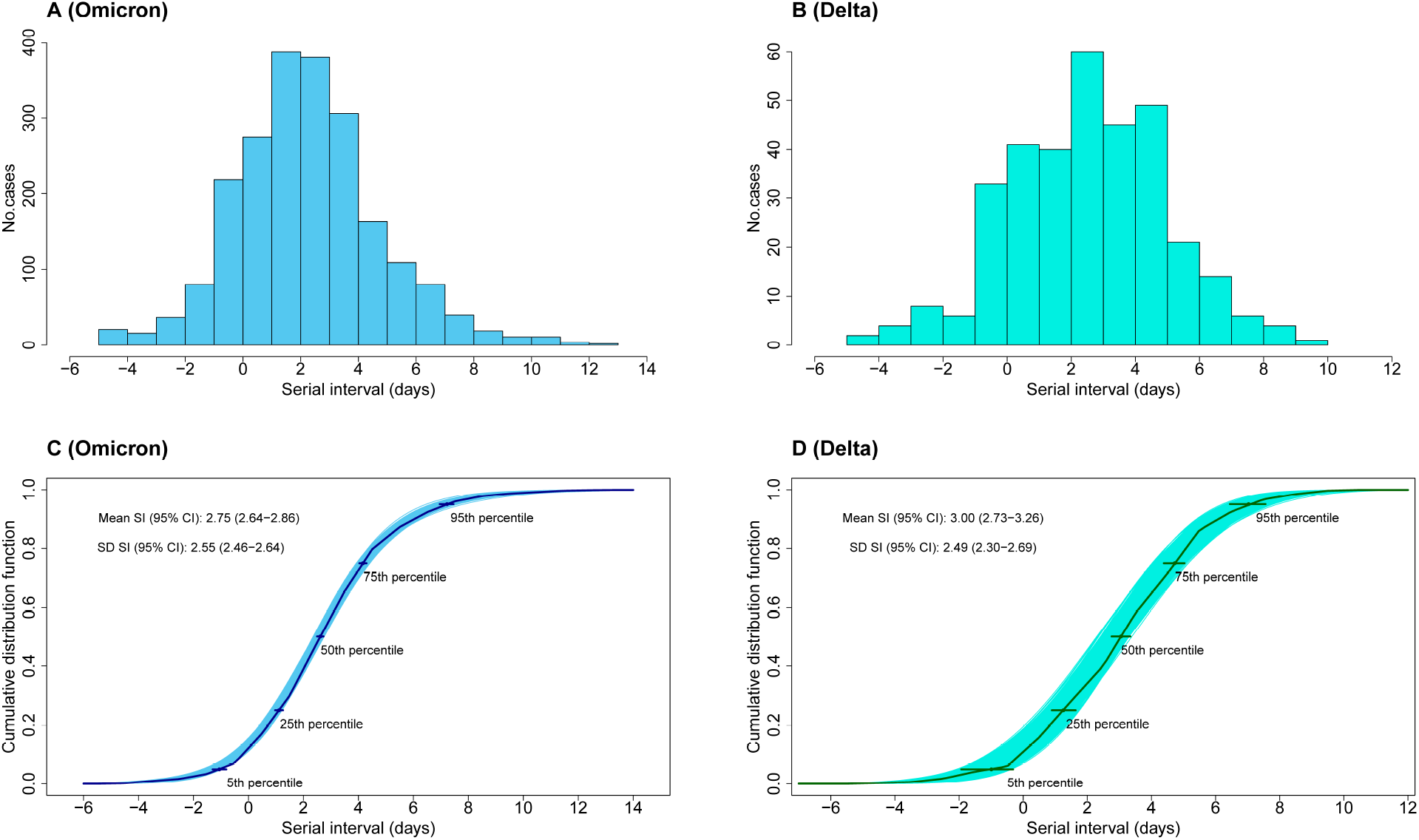
(**A**) Empirical distribution of serial intervals for SARS-CoV-2 Omicron. (**B**) Empirical distribution of serial intervals for SARS-CoV-2 Delta. (**C**) Nonparametric estimate 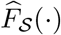 for Omicron (dark blue); smoothed estimates of the cdf for *B* = 5000 bootstrap samples (light blue) and 95% CIs for selected percentiles with associated point estimate (dark blue dot). (**D**) Nonparametric estimate 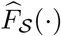 for Delta (dark green); smoothed estimates of the cdf for *B* = 5000 bootstrap samples (light green) and 95% CIs for selected percentiles with associated point estimate (dark green dot).

## 5 Conclusion

We propose a nonparametric approach to estimate the serial interval distribution of an infectious disease from illness onset data. The bootstrap technique is used to sample the nonparametric estimate of the cumulative distribution function and the generated samples can be used to compute point and interval estimates of any desired features of the serial interval. The proposed methodology has the following strengths and limitations.

### Strengths

Our method is entirely data-driven and does not require to input a parametric distribution for serial interval estimation. As such, we can directly sketch the main characteristics of the SI distribution without having to adjust parametric distributions to the data and compare which model fits best according to a given selection criterion (e.g. AIC, BIC or LOOIC). Also, if the modeler wants to fit a parametric distribution to the data, the nonparametric estimate of the cdf can be used as a benchmark to visually assess whether the chosen parametric model is in agreement with a data-driven fit, i.e. as an informal lack-of-fit test. Furthermore, our approach naturally deals with negative serial interval values. The bootstrap permits to compute interval estimates of any desired feature of *S*. Thus, confidence intervals are easily accessible and can be directly reported alongside point estimates following best practices outlined in Charniga et al. (2024). Algorithms underlying our nonparametric methodology are relatively simple and can be implemented at low computational cost. The small footprint of the associated code implies that it can be straightforwardly written in virtually any programming language most preferred by the user. The proposed method is available in the EpiLPS package Gressani (2021) and requires only minimal input by the user. Finally, the simple framework of our method favors reproducibility and facilitates serial interval analyses on past, current or future illness onset data streams.

### Limitations

For the moment, the proposed nonparametric method does not adjust for right truncation; a feature that may be encountered when serial interval data are observed in realtime. Another weakness of our approach is that the current bootstrap sampling process can only generate variates that are within the set of order statistics for the observed serial interval data. Methods exist to simulate variates beyond this range (see e.g. Kaczynski et al., 2012) and could be considered as a future improvement of our method.

## Data availability

Simulation results and real data applications in this paper can be reproduced with the code available on the GitHub repository (https://github.com/oswaldogressani/Serial_interval).

## Appendix

### Appendix A1

The bias, ESE and RMSE used in the simulation study of Section 3 to assess the performance of the point estimator of *θ*_*j*_ ∈ Θ_*S*_ are given by:

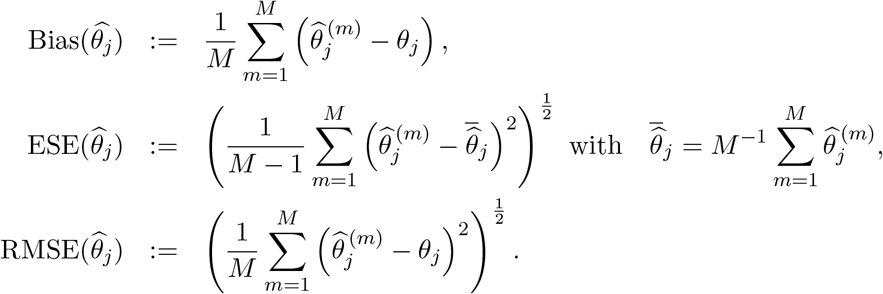

Performance of the interval estimator is measured through the coverage probability:

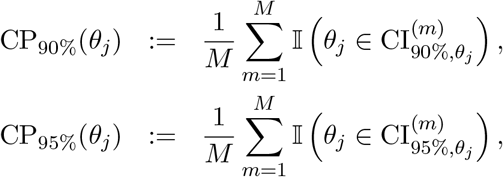

where 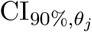 and 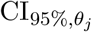 denote the 90% and 95% confidence interval, respectively, of *θ*_*j*_.

